# Predictive model of COVID-19 incidence and socioeconomic description of municipalities in Brazil

**DOI:** 10.1101/2020.06.28.20141952

**Authors:** Isadora C. R. Carneiro, Eloiza D. Ferreira, Janaina C. da Silva, Guilherme Soares, Daisy M. Strottmann, Guilherme F. Silveira

## Abstract

Coronaviruses are enveloped viruses that can cause respiratory, gastrointestinal, hepatic, and neurological diseases. In December 2019, a new highly contagious coronavirus termed severe acute respiratory syndrome coronavirus 2 (SARS-CoV-2) emerged in China. SARS-CoV-2 causes a potentially lethal human respiratory infection, COVID-19, that is associated with fever and cough and can progress to pneumonia and dyspnea in severe cases. Since the virus emerged, it has spread rapidly, reaching all continents around the world. A previous study has shown that, despite being the best alternative in the current pandemic context, social distancing measures alone may not be sufficient to prevent COVID-19 spread, and the overall impact of the virus is of great concern. The present study aims to describe the demographic and socioeconomic characteristics of 672 cities with cases of COVID-19, as well as to determine a predictive model for the number of cases. We analyzed data from cities with at least 1 reported case of COVID-19 until June 26, 2020. It was observed that cities with confirmed cases of the disease are present in all Brazilian states, affecting 36.5% of the municipalities in Rio de Janeiro State. The inhabitants in cities with reported cases of COVID-19 represent more than 73.1% of the Brazilian population. Stratifying the age groups of the inhabitants and accounting for the percentage of women and men does not affect COVID-19 incidence (confirmed cases/100,000 inhabitants). The demographic density, the MHDI and the per capita income of the municipalities with cases of COVID-19 do not affect disease incidence. In addition, if conditions are maintained, our model predicts 2,358,703 (2,172,930 to 2,544,477) cumulative cases on July 25, 2020.

## INTRODUCTION

A pandemic is defined as any epidemic disease widely distributed geographically that affects different regions simultaneously. Over the years, humanity has faced many instances when health and science are put to the test and need to present answers. The Spanish flu ravaged the whole world and was often confused with several other diseases, such as cholera, dengue, and typhus (GOULART, 2005). At the end of the pandemic, approximately 50 million people had died. In 2009, a novel influenza A (H1N1) virus emerged in Mexico, where the transmission remained geographically contained for at least one month. Migratory events and population movement caused the virus to spread to other countries, leading the World Health Organization (WHO) to declare H1N1 a global pandemic (LIPSITCH et al., 2011). In official WHO records, the H1N1 pandemic resulted in the deaths of 18,500 people around the world.

On March 11, 2020, after a declaration by the WHO, the world population again lived under a shadow of fear of a new pandemic, this time caused by a coronavirus. Coronaviruses (CoV) are enveloped viruses that are part of a large family of single-stranded RNA viruses with a positive-sense genome and can cause respiratory, gastrointestinal, hepatic, and neurological diseases. CoV can infect many animal species, including birds, cows, pigs, and humans, causing acute and chronic diseases (Chang et al., 2012; Weiss, 2011). Most infections caused by viruses from the family *Coronaviridae* induce a mild form of the disease in humans, usually causing flu-like symptoms. However, after the discovery of severe acute respiratory syndrome (SARS), a greater contagion capacity and lethality potential of this viral family was evidenced (Weiss et al., 2011). The etiological agent of SARS, SARS-CoV, was identified in mid-2003 after an outbreak of the disease in November 2002 in Guangdong Province, China, where 8,700 cases and 774 deaths were confirmed (Contini et al., 2020). The so-called novel CoV, initially referred to as 2019-nCoV, was first described when a group of patients reported symptoms of pneumonia of unknown cause in Wuhan City, Hubei Province, China, in December 2019 (ZHU, 2020).

On February 11, 2020, after phylogenetic and pathophysiological analyses, 2019-nCoV was officially named SARS-CoV-2 due to its similarity to SARS-CoV, as announced by the Coronavirus Study Group (CSG) of the International Committee on Taxonomy of Viruses (ICTV), according to the 2015 WHO nomenclature guidelines (Gorbalenya et al., 2020). The pathology caused by SARS-CoV-2 infection was termed COVID-19, characterized by a flu-like condition associated with fever and cough that can progress to pneumonia and dyspnea in more severe cases (CHAN et al., 2020). The incubation period of the disease varies from 2 to 14 days, and in approximately 80% of cases, infected individuals remain asymptomatic. However, unlike patients with influenza, viral transmission from asymptomatic individuals is possible (Contini et al., 2020). In addition, according to Contini (2020), the mechanism of contagion is direct, that is, through contact with respiratory fomites of infected people. Other studies show that SARS-CoV-2 can survive in the air for more than 3 hours and on surfaces such as plastics and metals for up to 3 days (Van Doremalen et al., 2020). Currently, there are no vaccines to prevent the disease, reinforcing the need for prophylactic measures, namely, correct hand, environment and surface hygiene and social distancing.

A previous study has shown that social distancing and other preventive measures alone may not be sufficient to prevent the spread of COVID-19, and the overall impact of the virus is of great concern (Sohrabi et al., 2020). It is also noteworthy that additional research is needed to help define the exact rates and mechanisms of person-to-person transmission, as well as to determine additional factors that can guide containment actions.

The internal and external logistic and transitory movements, as well as several other socioeconomic factors, can not only contribute to the understanding of viral spread but also assist in surveillance measures and competent decision-making for regional health systems, where such analysis can (and should) be implemented to reduce the exponential growth rate of positive cases. Different approaches are being used to better understand the transmission dynamics of SARS-CoV-2 to inform pandemic prevention and control measures. In this context, the present study aims to analyze the demographic and socioeconomic characteristics of cities with COVID-19 cases, as well as to adjust a predictive model for the cumulative number of disease cases thus expanding the possibilities of decision-making at the micro- and macroregional levels.

## MATERIALS AND METHODS

In the present work, an ecological study design was used; this method of epidemiological study helped us to generate hypotheses about possible associations between socioeconomic characteristics of the Brazilian municipalities and the COVID-19 incidence and fatality rate.

For the exploratory data analysis (EDA) and the predictive model adjustment, the Python programming language was used with several libraries specifically for this purpose. Along with Python, it was necessary to import different packages and libraries, the most used ones being pandas, NumPy and SciPy, with the function of organizing and structuring the data. For statistical calculations, the statsmodels package was imported to analyze the time series and the autoregressive integrated moving average (ARIMA) model for the forecasts. The purpose of these methods is to fit the model to the data as well as possible. Matplotlib and Seaborn were used to generate two-dimensional (2D) graphics. The project can be accessed through the GitHub page, https://github.com/gfsilveira/covid. The analyzed database contains data from 672 municipalities (12,1% of the cities in Brazil) in the 26 states of the federation plus the Federal District.

The records of COVID-19 cases at the municipal level were obtained through daily updates from the Health Departments of the Federative Units compiled by Álvaro Justen and his collaborators until June 26, 2020, available at https://brasil.io/dataset/covid19/caso. Demographic and socioeconomic characteristics publicly available at the municipal level, such as population density, Municipal Human Development Index (MHDI), total area in km^2^ and per capita income, were obtained from the Brazilian Institute of Geography and Statistics (IBGE) from the demographic census conducted in 2010. Data on the age range and sex distribution of the population were obtained from the 2015 census.

A data structure containing different age groups between 0 and 80+ years old from the IBGE 2015 census database was used. The age of residents in Brazilian municipalities was separated into groups of young (0 to 29 years), adults (30 to 64 years) and seniors (65+ years). This distribution grouped ages according to low risk (young), comorbidities risk (adults) and complications risk (seniors) from SARS-CoV-2. The percentage of declared sex data (female/male) was also analyzed in cities with cases of COVID-19. In addition to data from the population, the number of cities with COVID-19 cases within each state was analyzed by the percentage of Brazilian municipalities/states with confirmed cases until June 26, 2020.

Incidence (cases per 100,000 inhabitants) was analyzed according to demographic density (inhabitants/km^2^), Municipal Human Development Index (MHDI), socioeconomic data (per capita income), age groups and sex ratios.

For the development of the predictive model, we used the ARIMA model proposed by Box & Jenkins (1970), which consists of developing and adjusting stationary or nonstationary linear models relative to an observed time series. The autoregressive (AR) component indicates that the variable of the time series is regressed on its own lagged values. The I (for “integrated”) indicates that the data values have been replaced with the difference between n+1 and n values, performed more than once. The moving average (MA) is a calculation of data points by creating a series of averages of different subsets of full data. In the moving average, the regression error is a linear combination of various times in the past and error terms whose values occurred contemporaneously. The construction of the model was based on daily COVID-19 cases. To analyze the stationary condition of the time series in different orders, the augmented Dickey-Fuller (ADF) test was used. The null hypothesis of the ADF test is that the time series is nonstationary. The p-value of the test was 0.000234, on the order of differentiation 2, less than the significance level (0.05), rejecting the null hypothesis and indicating that the time series is indeed stationary (Figure 1G). To determine which predictive model to use, the autocorrelation function (ACF) (Figure 1B, E, H) and partial autocorrelation function (PACF) (Figure 1C, F, I) were analyzed to determine the ARIMA p, d, and q parameters. The ACF is the correlation of a variable with itself at differing time lags, and the PACF partial autocorrelation at lag 1 is very high (it equals the ACF at lag 1), but the other values are correlated when lag > 1. The PACF does not include any value for lag 0 because it is impossible to remove any intermediate autocorrelation between t and t−k when k=0, and therefore the PACF does not exist at lag 0. The ACF for an AR(p) process approaches zero very slowly, but the PACF goes to zero for values of lag > p. The ACF for an MA(q) process goes to zero for values of lag > q, but the PACF approaches zero very slowly. As defined by the ADF test, d = 2 was a second order of differentiation, making the series stationary (Figure 1G). In the second order of differentiation, the PACF with lag 2 is already below significance (Figure 1I), p = 1. The same occurs in the ACF in the second order, where lag 2 is below significance (Figure 1H). Then, the parameters were tested by the minor Akaike Information Criteria (AIC), four ARIMA models, (1,2,0), (1,2,1), (2,2,0), (2,2,1) and (2,2,2). With these data, the ARIMA parameters (2,2,1) showed the best adjustment (Figure 1J).

**Figure 1.**
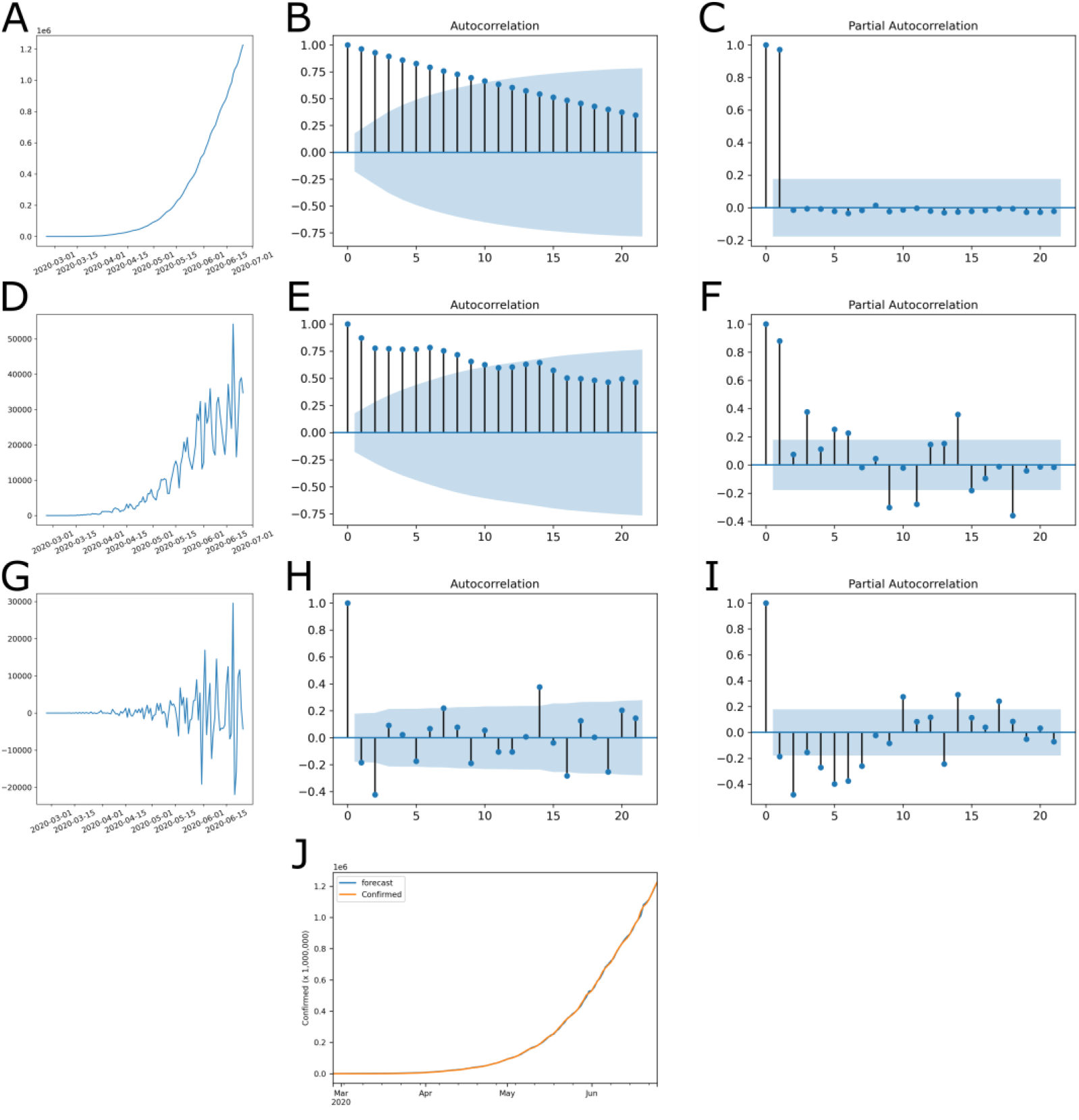
ACF and PACF plots for determination of model parameters. (A-C) Original series, (D-F) 1^st^-order differencing and (G-I) 2^nd^-order differencing. (A, D and G) Time series, with number of cases by date; (B, E and H) autocorrelation function; and (C, F and I) partial autocorrelation function. With this data, a model ARIMA (p=1, d=2, q=1) was determined. (J) Adjustment of forecast data with observed outcome data.

## RESULTS

### 1) A total of 73.1% of the Brazilian population lives in cities with confirmed cases of COVID-19

Since COVID-19 is a pathology caused by SARS-CoV-2 that is transmitted directly from person to person, in the present work, we seek to observe the characteristics of the affected cities. The analyzed database contains data from 672 municipalities (12,1% of the 5,570 cities in Brazil) from the 26 states of the federation, plus the Federal District (Figure 2), which had at least 1 confirmed infection as of June 26, 2020, totaling 1,225,993 (0.58% of the Brazilian population) cases of COVID-19, which resulted in 54,918 deaths. The most affected state was Rio de Janeiro, reaching 36.5% of the municipalities reported cases of COVID-19. Until the last analyzed date, we observed an incidence rate of 63.31/100,000 inhabitants and a mortality rate of 2575 (per 100,000 inhabitants). The data analyzed in this work have been updated daily since February 25, 2020. The first 100 cases had been confirmed by March 14, and after one week, on March 21, there were already 1,000 confirmed cases. Brazil reached 10,000 and 100,000 confirmed cases on April 4 and May 3, respectively. As of Jun 19, the number of confirmed cases reached 1,039,339.

**Figure 2.**
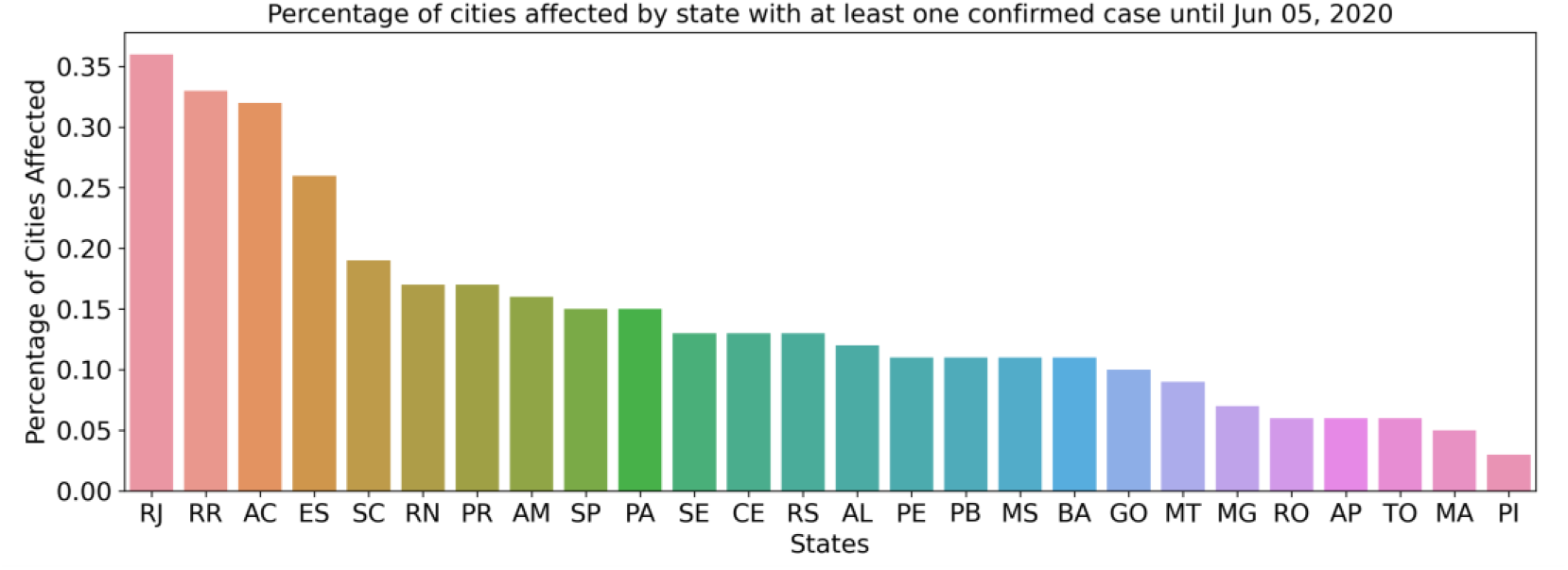
All Brazilian states have confirmed cases of COVID-19. Distribution of confirmed cases of COVID-19 by state. Percentage of cities with at least 1 case of positive infection.

The population of Brazil in 2018 (last available data) was 210,147,125 inhabitants, with the largest number of inhabitants between 10 and 34 years old (Figure 3A). The cities where cases of COVID-19 were observed have 153,528,953 inhabitants, representing 73.1% of the Brazilian population, and as expected, the distribution of age groups is the same as the general distribution in Brazil (Figure 3B). In the present study, the age ranges of the population in the affected cities were grouped into seniors over age 65, with a higher risk; adults between 30 and 64 with a greater likelihood of comorbidity; and the young, aged 0 to 29, with lower risk. The percentage of inhabitants in each age group (Figure 3C) was be used for the analysis in relation to COVID-19 incidence. The city with the greatest incidence, Santo Antônio do Içá (Amazonas) (4592.17 cases/100,000 inhabitants), has 70% of inhabitants in the young group (0-29 years old), 27% in the adult group (30-64 years old) and 3% in the seniors group (65+ years old). In contrast, the city of Utinga (Bahia), with a lower COVID-19 incidence in the database (15,64 cases/100,000 inhabitants), has 55% of inhabitants in the young group (0-29 years old), 38% in the adult group (30-64 years old) and 7% in the seniors group (65+ years old). The quartiles of the percentage of inhabitants in each age group were analyzed to determine if cities with a large percentage of senior, adult or young people have a greater incidence. For the young group, the percentage of inhabitants was divided into 34 – 44% (Figure 3D, blue), 45 – 48% (Figure 3D, orange), 49 – 51% (Figure 3D, green), and 52 – 71% (Figure 3D, red) of the total population. The adult group composed 26 – 41% (Figure 3E, blue), 42 – 45% (Figure 3E, orange), 46% (Figure 3E, green), and 47 – 52% (Figure 3E, red) of the total population. The senior group included 2 – 6% (Figure 3F, blue), 7 – 8% (Figure 3F, orange), 9 – 10% (Figure 3F, green), and 11 – 16% (Figure 3F, red) of the total population. These groups were analyzed against the incidence of COVID-19 (Figure 3D-F). In conclusion, in the affected cities, the age groups do not show a relationship with the incidence of the disease. Municipalities with a greater or lesser percentage of inhabitants in the young, adult, or senior age groups do not have a greater or lesser incidence of COVID-19.

**Figure 3.**
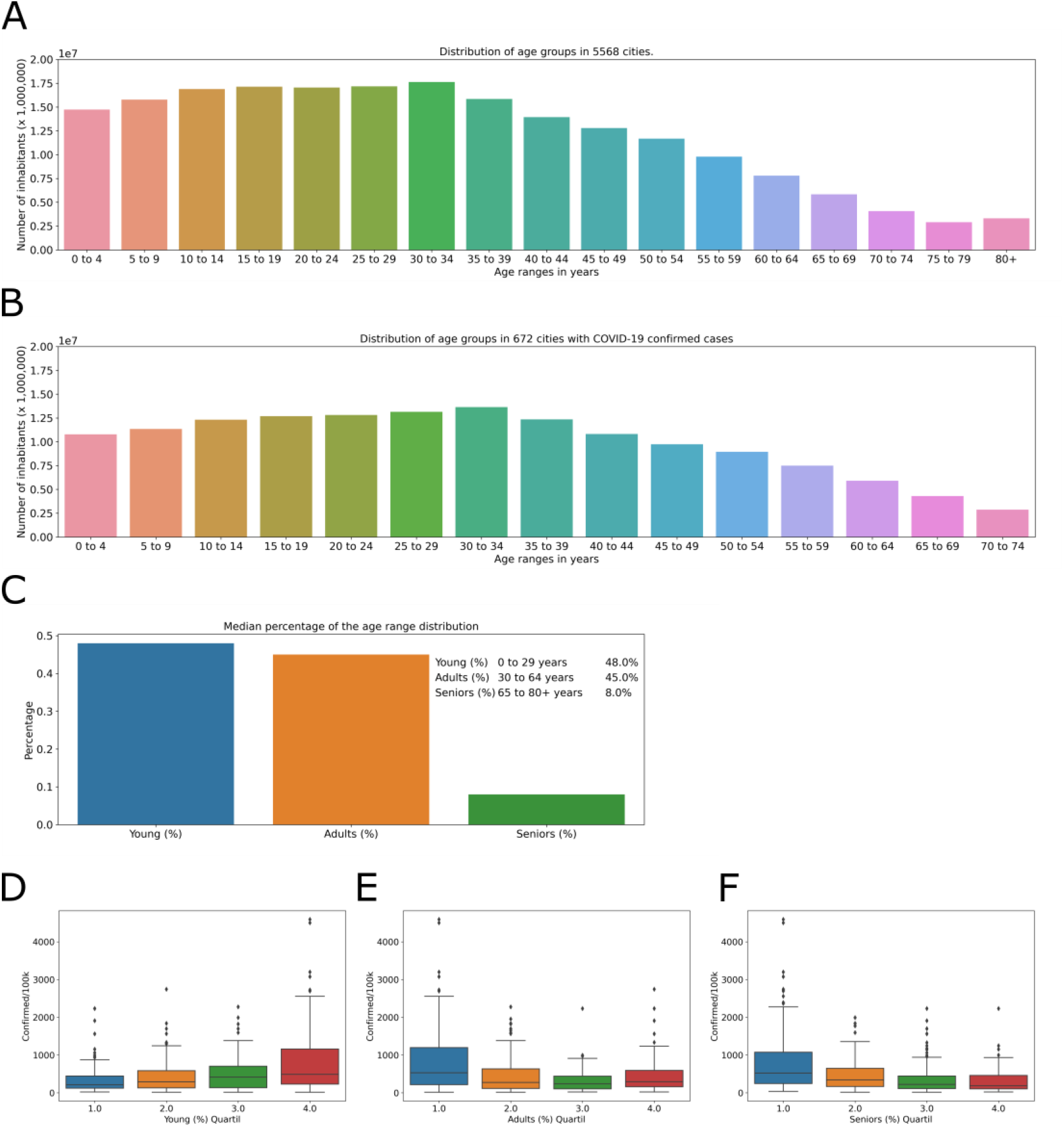
Age distribution in cities with cases of COVID-19. (A) Number of inhabitants by age group (increments of 5 years) in Brazil. (B) Number of inhabitants by age group in cities with cases of COVID-19. Stratification of age groups by age in the municipalities studied, represented by the percentage of the number of inhabitants for each city (C). Distribution of quartiles of the number of inhabitants per age groups of (D) young, (E) adults, (F) seniors, in relation to the COVID-19 incidence, in the affected cities.

Another demographic factor analyzed was the declared sex of the inhabitants. In Brazil, the population distribution percentage is 51.7% for women and 48.3% for men. We observed a similar distribution, with 50.11% (2,979,950) women and 49.89% (2,965,800) men, in cities with COVID-19 cases. The quartiles of the number of women and men inhabitants in the affected cities do not show a relationship with the incidence of the disease. For the male group, 46 – 49% (Figure 4A, blue), 50% (Figure 4A orange), 51% (Figure 4A green), and 52 – 69% (Figure 4A red) of the total population were included. For the female group, the percentage of inhabitants was divided into 31 – 49% (Figure 4B, blue), 50% (Figure 4B, orange), 51% (Figure 4B, green), and 52 – 54% (Figure 4B, red) of the total population. Municipalities with a greater or lesser number of men (Figure 4A) or women (Figure 4B) inhabitants do not have a greater or lesser incidence.

**Figure 4.**
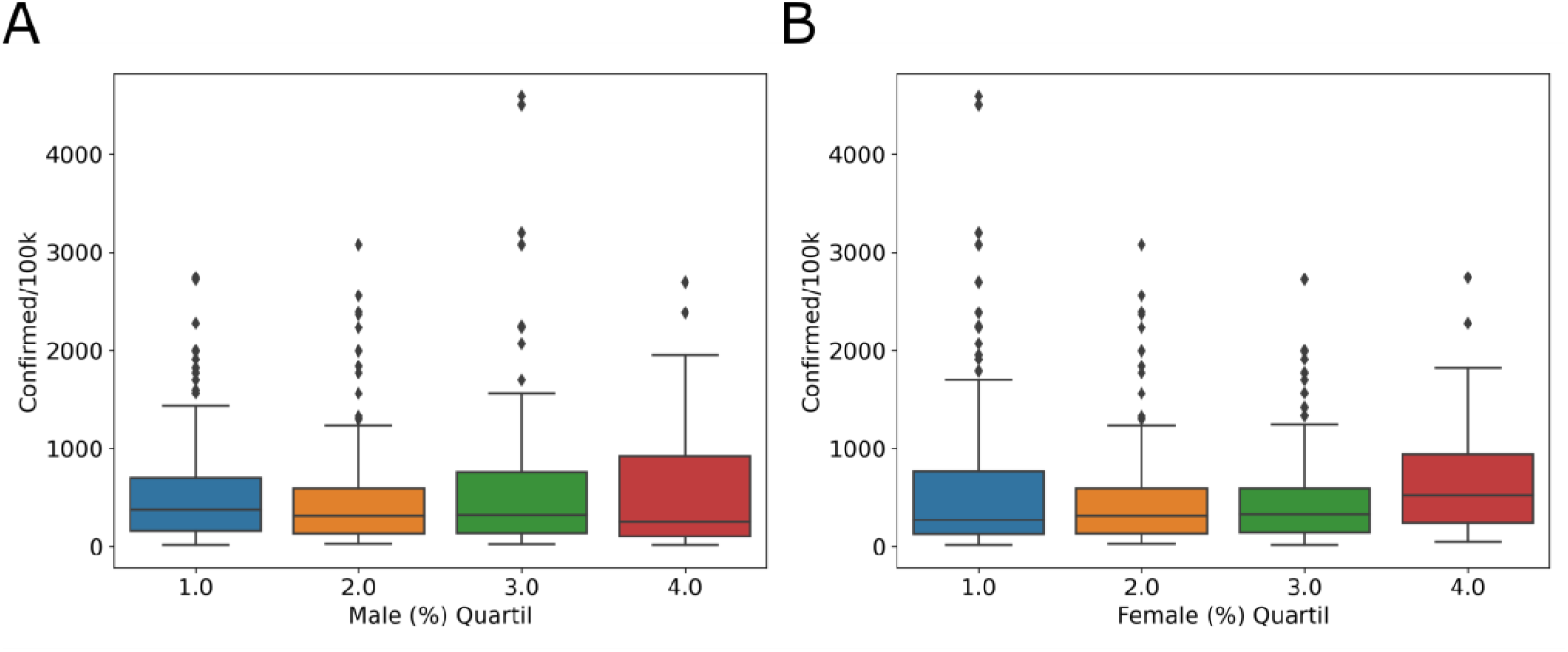
Declared sex of inhabitants in cities with cases of COVID-19. Distribution of quartiles of the percentage of (A) male and (B) female inhabitants, in relation to the COVID-19 incidence, in the affected cities.

Since SARS-CoV-2 is transmitted from person to person, we describe characteristics, such as demographic density (inhabitants/km^2^), of the dispersion and agglomeration of people in the municipalities. Of the 672 cities analyzed, 577 (86%) are more densely populated than the national average density, 23.9 hab/km^2^. In the database, the mean population density was 683.76 hab/km^2^ in all cities with COVID-19 cases. The city with the lowest demographic density was Novo Airão (Amazonas), with 0.39 hab/km^2^ and 1074.33 cases/100,000 inhabitants. The highest demographic density was in São João de Meriti (Rio de Janeiro), with 13024.6 hab/km^2^ and 334.881 cases/100,000 inhabitants. These apparent differences have no significance in the grouped data. The COVID-19 incidence was observed for the demographic density (hab/km^2^) quantiles, (Figure 5A) 0.39 to 38.86; (Figure 5B) 38.86 to 127.27; (Figure 5C) 127.27 to 540.95; and (Figure 5D) 540.95 to 13024.56. The analyses of the quartiles of demographic density show no relation with COVID-19 incidence (Figure 5E). Municipalities with a greater or lesser demographic density do not have a greater or lesser incidence of COVID-19.

**Figure 5.**
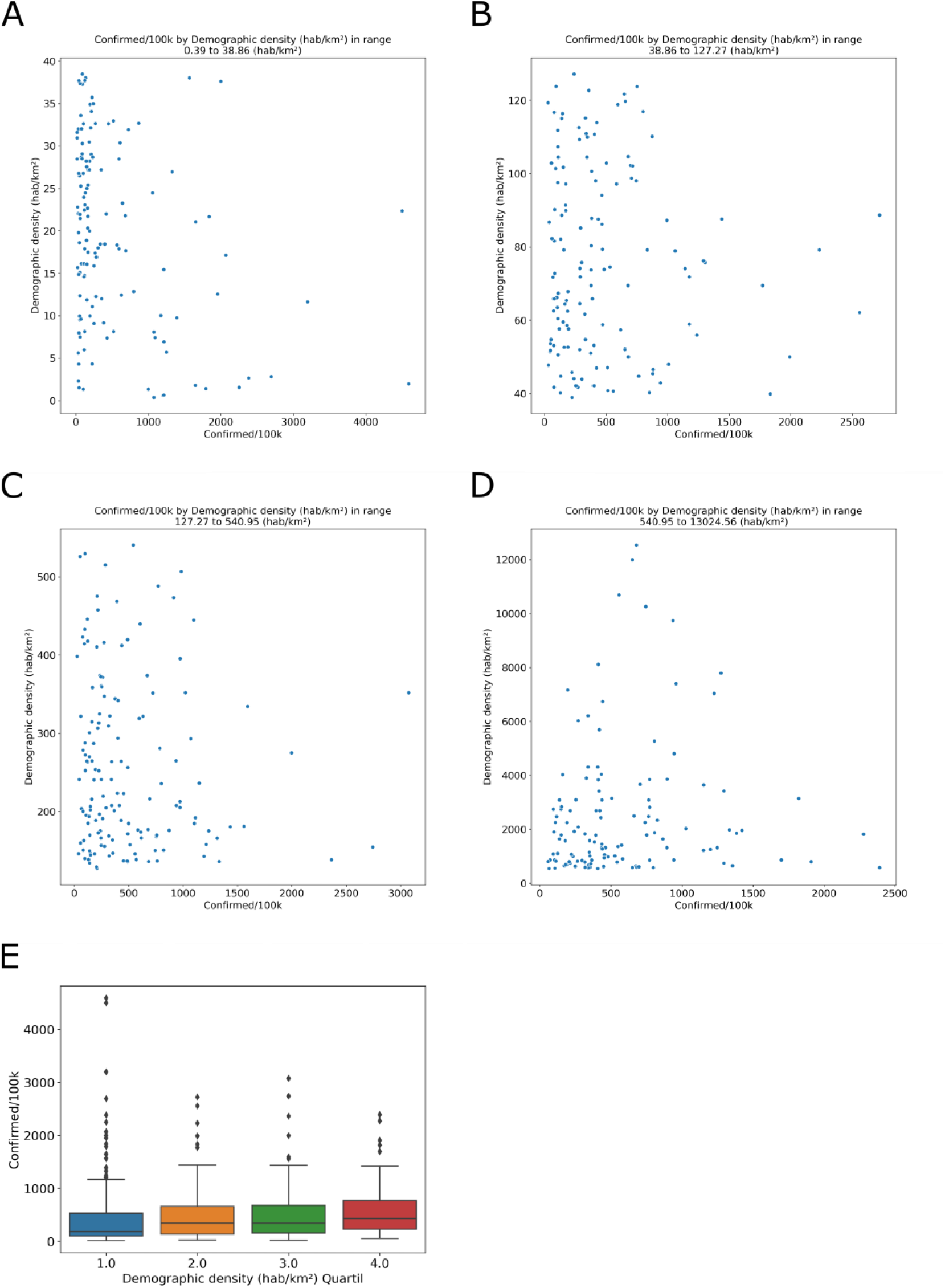
The demographic density in cities with cases of COVID-19. Number of inhabitants (population) by area (km^2^) resulting in demographic density (hab/km^2^) in four quantiles, (A) 0.39 to 38.86; (B) 38.86 to 127.27; (C) 127.27 to 540.95; and (D) 540.95 to 13024.56. (E) The relationship between the demographic density quartiles and COVID-19 incidence.

The Municipal Human Development Index (MHDI) is related to life expectancy, educational level and income distribution. The global Brazilian HDI for 2013 was 0.744, the 79th position in the world, ranking among the 187 countries and territories recognized by the United Nations. In the Global HDI for HDR 2014, the three dimensions have the same weight, and the human development ranges are fixed as follows: low human development, less than 0.550; average, between 0.550 and 0.699; high, between 0.700 and 0.799; and very high, above 0.800. In Brazil, the per capita income was 1.48 (R$ 1,443.10) for 2017 (last year with available data). In this context, the MHDI (Figure 6A) and per capita income (Figure 6B) were compared with COVID-19 incidence.

**Figure 6.**
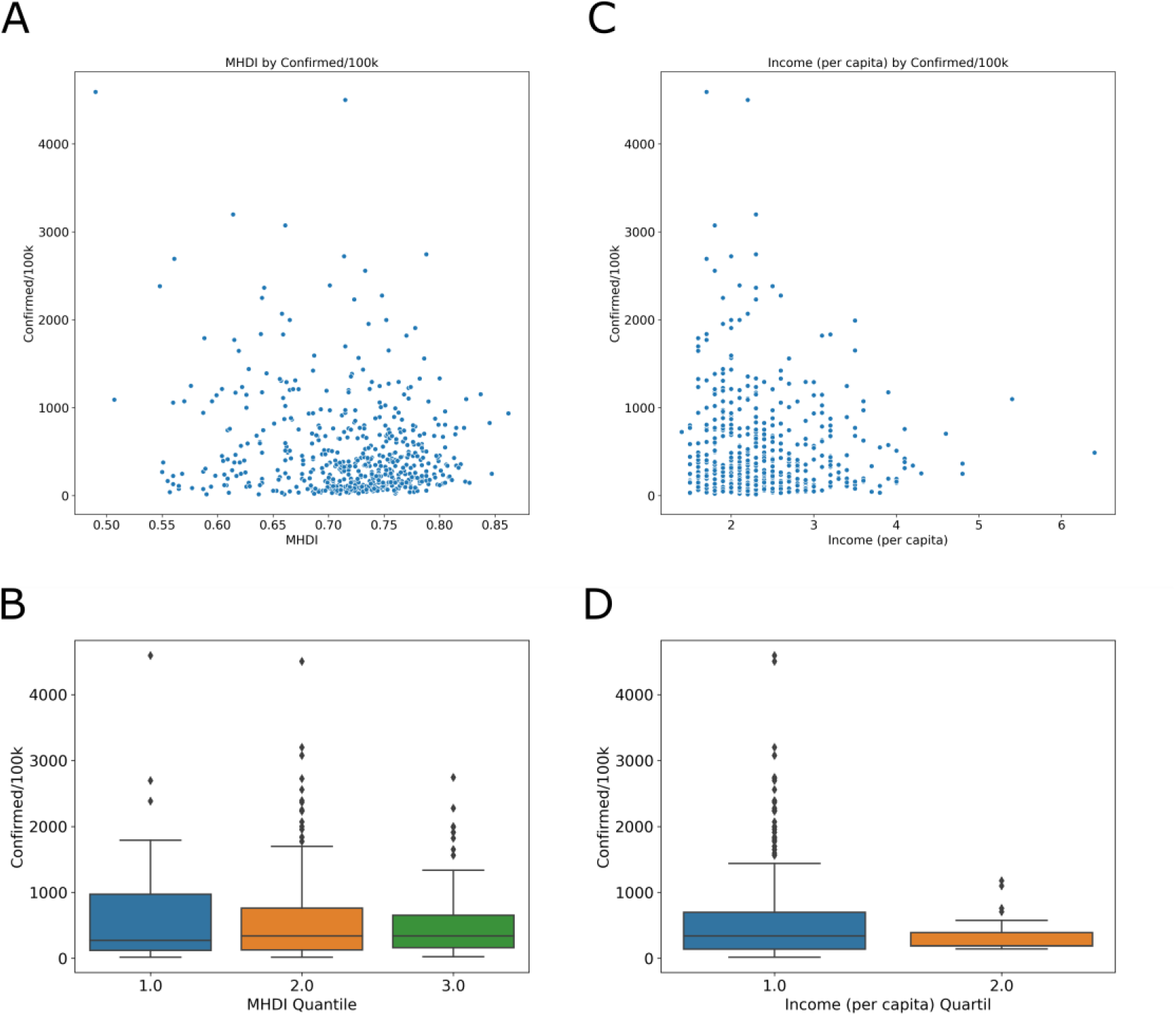
The Municipal Human Development Index (MHDI) and per capita income in cities with cases of COVID-19. (A) MHDI and (C) income (number of times the minimum wage [R$ 975.00] is earned per month for formal workers) in cities with cases of COVID-19. (B) The relationship between the MHDI-low 0.49 to 0.613 (blue), MHDI-mean 0.614 to 0.737 (orange) and MHDI-high 0.738 to 0.862 (green) groups. (D) The income was divided into low 1.4 to 3.8 (blue) and high 3.9 to 6.4 (orange).

The results show that most cities with positive cases for COVID-19 are above the national average for both MHDI and per capita income. With the goal to observe the relationships in the data sets in greater detail, we analyzed the MHDI divided into low 0.49 to 0.613 (Figure 6B, blue), mean 0.614 to 0.737 (Figure 6B, orange) and high 0.738 to 0.862 (Figure 6B, green) groups. The income was divided into low 1.4 to 3.8 (Figure 6B, blue) and high 3.9 to 6.4 (Figure 6B, orange). The socioeconomic index does not show a relationship with the incidence of the disease.

Once some of the characteristics of the cities with cases of COVID-19 were described, we sought to determine a model for predicting infection using the time series of confirmed cases.

#### 2) For July 25, 2020, the evolution model predicts 2,358,703 (2,172,930 to 2,544,477) confirmed cases

Due to the current level of infection in the cities analyzed, the scarcity of data does not allow the development of a robust predictive model for cases confirmed at the municipal level. To understand the condition of the infection at the national level, we analyzed the time series of accumulated data for confirmed cases. There was a clear upward trend in the number of cases (data not shown). To suggest a prediction for the evolution of COVID-19 cases in Brazil, we use computational modeling in the time series. The best adjusted model for the forecast was ARIMA(2,2,1) using data from the last 30 days, which forecasts 2,358,703 cumulative cases on July 25, 2020, with a 95% confidence interval of 2,544,477 to 2,172,930 (Figure 7).

**Figure 7.**
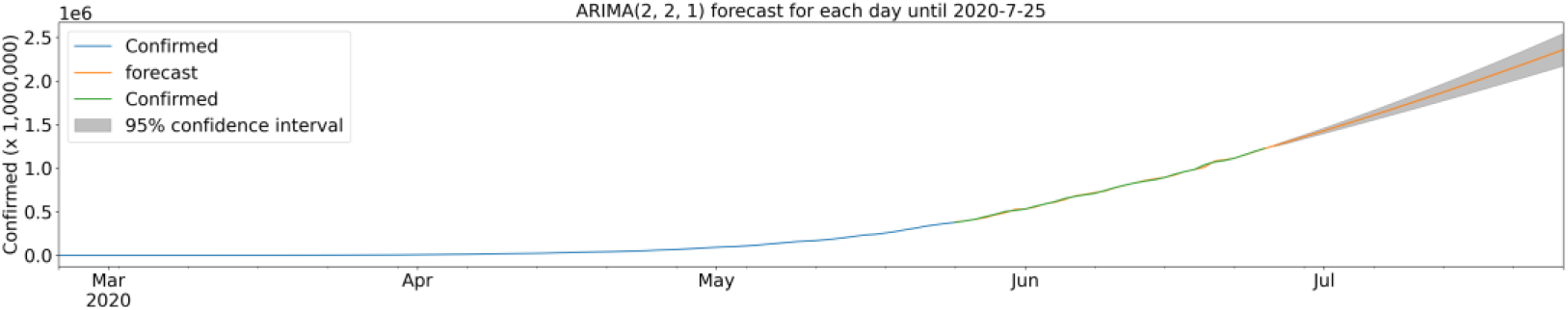
Average estimate of 2,358,703 cumulative confirmed cases in 30 days. ARIMA model of the forecast of confirmed cases until July 25, 2020. Confirmed cases (blue), forecast (orange), model fit analysis (green) and forecast with 95% confidence interval (gray). Up to the end date, between 2,544,477 and 2,172,930 cases are expected.

## CONCLUSION

Altogether, 672 cities, accounting for 73.1% of the Brazilian population, had at least 1 case of COVID-19 by June 26, 2020. The age distribution of the inhabitants in those cities, which include the most populous cities of Brazil, remains the same as the average age distribution of the Brazilian population. The average distribution of women and men in the cities studied also corresponds to the national average. The demographic density, the MHDI and the per capita income of the municipalities with cases of COVID-19 are above the national average. However, there seems to be no relationship between the indexes analyzed and the incidence of COVID-19 in these cities, suggesting that other factors (virulence, immune background) may influence the spread of the disease. Our model predicts 2,358,703 cumulative cases (2,172,930 to 2,544,477) on July 25, 2020.

## DISCUSSION

In this work, we studied the recent occurrence of COVID-19, a respiratory disease caused by the coronavirus, SARS-CoV-2, which originated in the city of Wuhan, China, and analyzed the correlation of transmission and death rates, through confirmed cases using Brazilian demographic and socioeconomic data. Knowledge of the demographic distribution and socioeconomic situation of the population becomes significant when comparing transmission and death rates across the country.

Brazil currently has 5,570 municipalities in 26 states distributed disproportionately over a total area of 8,511,000 km^2^ (IBGE, 2019). As described in the results section, municipalities with a population larger than 295,955 inhabitants showed a positive correlation between the size of the population and the number of confirmed cases of the disease. However, of all the municipalities, only 95 have a population of over 295,955 inhabitants, including all state capitals and the Federal District. The 95 municipalities most populous represent 1.71% of the country’s total cities, with the majority, 94.22% (5,245 municipalities), of Brazilian cities having a population less than or equal to 100,000 inhabitants (IBGE, 2019).

Therefore, based on our results, it is possible to assert that the transmission of the disease is more likely to impact less than 2% of Brazilian municipalities. However, it is essential to reinforce that the 95 most populous cities in the country, together, are home to 83,951,535 inhabitants, which represents 40% of the total population of Brazil, with a current demographic density of 205.5 million people (IBGE, 2018). Thus, neglecting the recommendations of the WHO about isolation and social conduct, in a time of a pandemic, is an attack on public health in Brazil.

When analyzing the data in relation to the states of the federation, we noted similar observations as those obtained in relation to the municipalities. The state of São Paulo is the most populous in the country and in turn is the state with the highest number of confirmed cases of COVID-19, with 11,043 cases confirmed to date, according to the Ministry of Health (2020). It is important to note that the actual numbers of cases and deaths from the disease may be different from the official data, taking into account the impact that the delay in reporting has on the estimates and that reported cases depend on hospitalization (FIOCRUZ, 2020).

When we take into account the data obtained, it is possible to show that social isolation is a valid measure to be applied in municipalities that have a resident population larger than 295,000 inhabitants. For these municipalities, the more intense the measures, the flatter the transmission curve becomes; thus, hospitals and health units can have greater control of the situation under the demand of patients who require specialized care.

However, it is important to mention that it is not possible to conclude the real importance of social isolation in municipalities with a population below the aforementioned number; however, according to the results, there is a negative correlation between demographic density and the number of cases in these cities, which, in theory, would indicate a lack of connection between these aspects.

According to Hellewell et al. (2012), in a study to evaluate measures to contain the transmission of the disease, social isolation is insufficient to control the outbreak, requiring new interventions to achieve control of the transmission of the disease. However, isolation can contribute to spreading the overall size of an outbreak over a longer period of time (Hellewell et al., 2012). Taking this into account, it becomes possible to assess the importance of measures of social isolation, even for municipalities with a small population, demonstrating the great importance of such measures that should be intensified in the most populous cities and not neglected in cities with fewer than 295,000 inhabitants. Therefore, it is interesting to evaluate the average traffic of the Brazilian population mainly in the forms of essential workers, e.g., truck drivers, who supply basic necessities. This supply is primarily carried out in Brazil through land transportation, often long distance, between the capitals and other municipalities of the federation. During the month of March, an average of 1000 trucks arrived at the Supply Center of the Federal District every Monday and Thursday (CEASA, 2020).

It would be interesting, as a future perspective, to further these studies in order to develop a system of equations that could indicate a proportional factor of the relationship between population and lethality rate to compare the fold-change in the lethality in a more populous city to that of a less populous city. Perhaps, as the number of inhabitants in a city doubles or triples, lethality does not necessarily double or triple, indicating that there is no linearity in the cases.

Therefore, the information reported in this study allows us to highlight that cities with a higher number of inhabitants who choose not to comply with social isolation have a higher risk and probability of infection. However, it is essential to show that infection with SARS-CoV-2 is not due to the simple fact of living in a more or less populous city and that there are different regional characteristics, both geographic and socioeconomic, that can influence dispersion, not only for SARS-CoV-2 but also for many other pathogens (Mogi and Spijker, 2020; Dowd et al., 2020).

Thus, it is worth emphasizing once again that in the current pandemic context, without effective prophylactic and therapeutic treatments, social isolation has proven to be an efficient measure to control outbreaks in the most populous cities. Therefore, our findings suggest that this approach should indeed not just be simulated but also applied to reduce transmission and avoid hospital demands above service capacity, to provide care for all patients at the micro- and macroregional levels (FIOCRUZ, 2020).

## Data Availability

The project can be accessed through the GitHub page.

https://github.com/gfsilveira/covid

## ACKNOWLEDGMENTS

We would like to thank Hellen Geremias dos Santos, PhD, for the critical reading of the work. We would like to thank the Creative Commons Attribution ShareAlike and Álvaro Justen and collaborators from Brazil. We thank IO for the confirmed case database. We would like to thank the scholarship funding agencies CAPES and Fiocruz.

